# Swiss Digital Pathology Recommendations: Results from a Delphi process conducted by the Swiss Digital Pathology Consortium of the Swiss Society of Pathology

**DOI:** 10.1101/2023.09.15.23295616

**Authors:** the Swiss Digital Pathology Consortium, Andrew Janowczyk, Inti Zlobec, Cedric Walker, Sabina Berezowska, Viola Huschauer, Marianne Tinguely, Joel Kupferschmid, Thomas Mallet, Doron Merkler, Mario Kreutzfeldt, Radivoje Gasic, Tilman T. Rau, Luca Mazzucchelli, Isagard Eyberg, Gieri Cathomas, Kirsten D. Mertz, Viktor H. Koelzer, Davide Soldini, Wolfram Jochum, Matthias Rössle, Maurice Henkel, Rainer Grobholz

## Abstract

Integration of digital pathology (DP) into clinical diagnostic workflows is increasingly receiving attention as new hardware and software become available. To facilitate the adoption of DP, the Swiss Digital Pathology Consortium (SDiPath) organized a Delphi process to produce a series of recommendations for DP integration within Swiss clinical environments. This process saw the creation of 4 working groups, focusing on the various components of a DP system (1) Scanners, Quality Assurance and Validation of Scans, (2) Integration of WSI-scanners and DP systems into the Pathology Laboratory Information System, (3) Digital Workflow – compliance with general quality guidelines, and (4) Image analysis (IA)/artificial intelligence (AI), with topic experts for each recruited for discussion and statement generation. The work product of the Delphi process is 83 consensus statements presented here, forming the basis for “SDiPath Recommendations for Digital Pathology”. They represent an up-to-date resource for national and international hospitals, researchers, device manufacturers, algorithm developers, and all supporting fields, with the intent of providing expectations and best practices to help ensure safe and efficient DP usage.

## Introduction

Clinical Pathology is in the process of undergoing a digital transformation, wherein routinely produced glass slides are no longer read in an “analog” manner using a microscope but are instead viewed in a “digital” manner on computer screens after digitization. This digital pathology (DP) paradigm offers a number of important advantages, many of which are now being realized in clinical routines^1^. Improvements in streamlining of pathology practices, workflows, and quality of life enhancements for pathologists have already been seen. For example, DP streamlines pathology practices by having pathologists access and analyze slides remotely, eliminating the need for the organization, and physical transportation, of glass slides^2^. DP further facilitates collaboration among pathologists, enabling them to easily share and discuss cases, which can lead to more accurate and timely diagnoses^3^. Moreover, DP is staged to reduce costs associated with slide storage and management^4^, as digital images can be stored electronically and accessed when needed, potentially eliminating the need for long-term physical archives. These same repositories provide rapid retrieval of previous cases that may be of comparative interest.

Beyond these clinical improvements, substantial work is demonstrating that once these images are digitized they can be employed by computational approaches geared towards predicting diagnosis, prognosis, and therapy response of patients^5^. These DP tools and image based biomarkers leverage the present day confluence of 4 factors for their success: (a) relatively inexpensive computational power in the form of graphics processing units (GPU), (b) inexpensive storage of the large file sizes typically associated with DP, which can often reach more than 2GB per slide, (c) increased generation of whole slide images (WSI) via adoption of digital slide scanners in both research and clinical use cases, with some institutions routinely producing more than 2,000 whole slide images *per day*, and (d) new algorithms, such as deep learning^6^, whose success continues to be built upon the availability of the other factors.

In contrast, disadvantages associated with DP appear to be connected with its setup and instantiation, as opposed to long-term sustainment and usage. Challenges associated with initial cost^7^, software and hardware integrations, refinement of lab practices^8^, and additional training requirements potentially disrupting workflow and productivity are not uncommon during early stages of DP deployment. Secondary issues, associated with e.g., slide scanning time, standardization of work product, and compliance with regulatory and legal issues are likely connected to lack of experience and detailed planning, thus benefiting from and motivating the need for sharing of points for consideration and best practices.

In spite of any limitations, the opportunities afforded by going digital appear to be driving substantial investments by academic researchers, hospitals, and industry to put in place validated DP workflows for clinical usage^9^. As a result, groups of motivated experts have been formed both nationally and internationally to engage in knowledge sharing and best practices. For example, our Swiss Digital Pathology Consortium (SDiPath) was founded as a working group of the Swiss Society of Pathology (SSPath) in 2016 and now enlists over 170 members, evenly split between pathologists (defined here in the broadest sense, including board-certified pathologists, neuropathologists, dermatopathologists, residents, and trainees), computational pathology researchers, and technical experts which enable DP activities (i.e., histology-technicians and information technology specialists).

A common theme emerging from the development of our own vision for a national DP infrastructure^10^, to surveys regarding DP usage and adoption^11,12^, is the apparent need for national recommendations for the deployment and validation of DP pipelines, workflows, and algorithms. This is in line with efforts in other countries and organizations that have produced similar recommendation documents, geared towards their specific needs and regulatory environments (e.g., Germany^13^, Australia^14^, United States of America^15^, United Kingdom^16^). These efforts express the importance associated with producing consistent work product, documenting workflows, estimating both human and technological costs; together serving the tenet of patient safety having paramount importance. Notably, it is a requirement that the digital transformation of DP should not yield inferior performance, safety, or quality assurances as compared to its microscope based analog counterpart, as reflected by respective CE, FDA, or IVD certifications.

Given the nascent nature of clinical DP instantiation, and the associated cross-domain skillset needed, a concerted effort of agglomerating different stakeholders’ experiences and opinions is warranted. This is especially the case as digital workflows are often non-trivial to materialize, and may further be burdensome to upgrade or rectify if unexpected issues arise^17,18^. There are often unforeseen challenges, for example, those associated with incorrect scope definition as discussed in our previous work entitled “Going digital: more than just a scanner!”^17^. While claims that “Digital Pathology: The time has come”^18^ are emerging, there appears to remain potential hesitancy to engage in a digital transformation without clear guidelines of expectations and deliverables^11,12^. The recommendations presented here, similar to those produced in other countries, employed surveys and discussions with experts in their respective fields to curate experiences and thoughts. The ultimate goal is not only to provide best-practices and suggestions to those at different stages of their digital transformation, but to take current potentially ad-hoc approaches and solidify them into common practices to the benefit of pathologists, regulators, device and algorithm manufacturers, researchers, and above all, our patients.

## Methods

To build consensus on a set of DP recommendations from SDiPath members, a Delphi Process was used. Briefly, this process consists of rounds wherein (1) participants vote on their level of agreement with provided statements, (2) discordant statements are reviewed, discussed, and revised, and (3) new statements are submitted for voting again until a consensus is reached.

To facilitate this process, four working groups were formed around major pillars associated with DP: (1) Scanners, Quality Assurance and Validation of Scans, (2) Integration of WSI-scanners and DP systems into the Pathology Laboratory Information System, (3) Digital Workflow – compliance with general quality guidelines, and (4) Image analysis (IA)/artificial intelligence (AI). These working groups were led by experts in their respective areas, who were tasked with recruiting members to their WGs having relevant expertise as needed to generate a series of statements. On average, each working group consisted of approximately 10 people. Working groups were encouraged to review existing guidelines from other organizations, such as the Digital Pathology Association^19^, CAP^15^, Canadian^20^, UK^16^, German^13^, Korean^21^, and Australian^14^ guidelines^22^, and use them to critically reflect on their own statements.

Nomenclature was suggested such to indicate level of severity of proposed statements, with (a) ‘must’ indicating an imperative, (b) ‘should’ indicating a suggestion, and (c) ‘could’ indicating preferable but not required.

After the individual working groups formulated their statements, they were unified into a single document, in which all working group members reviewed and provided feedback. In total, 83 statements were created and voted upon at a WG level via Google Forms, such that there was 1 form per WG, to allow participants to selectively engage with WG’s matching their expertise. Participants were asked to select between: (i) Strongly Agree, (ii) Agree, (iii), Neutral, (iv) Disagree, and (v) Strongly Disagree for each statement. The demographics of the expertise and background of the participants was recorded and is provided below. The survey was announced via various venues including the SDiPath mailing list, in person meetings, and direct departmental level recruitment.

After a 1 month waiting period for feedback, from between May 2022 and June 2022, 14 statements were identified as needing discussion and clarifying language at the WG level. These statements were returned to the working groups wherein they underwent a supervised revision with the experts to modify the statements based on comments provided by the voting members. These were then again reviewed by all working groups for approval before being submitted to the members for a second round of Delphi voting via a single unified Google Form. This round was made available in November 2022 for 2 weeks, after which a review of the participant votes and feedback indicated convergence. Importantly, the voting members represented a diverse set of Swiss pathology stakeholders, hailing from all over the country. Those pathologists involved in the production of these guidelines are affiliated with all 5 University Hospitals, as well as 4 Cantonal Hospitals, and 2 Private Institutions.

Statement responses are reported in descending percentage order. Consensus was determined as being reached if >66% of all voters “Agreed” or “Strongly Agreed”. All of the statements presented here reached that level of agreement, indicating full consensus.

Participation was entirely voluntary, and there was no financial compensation for study participation and no disadvantage related to non-participation.

## Results

### Working Group 1 – Scanners, Quality Assurance and validation of Scans

This Working Group focuses on scanners, quality assurance, and the validation of scans in digital pathology.

These recommendations emphasize the importance of clear workflow definition, scanner evaluation, and thorough validation processes in digital pathology. These statements were asked to focus on the first part of the digital pathology pipeline – the selection, installation, and validation of whole slide image scanners. They discuss workflow creation and adjustment, documentation requirements, ideal scanner properties, and approaches for scanner validation. The recommendations are summarized as follows, with individual statements and agreement levels provided in Appendix A.

### Scope of the Diagnostic Workflow of Digital Pathology

- Define the scope of the targeted digital pathology workflow, considering different types of workflows (e.g., diagnostic biopsy, special stain, image analysis).
- Create a well-documented standard operating procedure (SOP) that describes the entire workflow, including scanning, and is in line with quality management systems and accreditation requirements.
- Establish security settings for authorized access to workflow components.
- Prioritize workflows based on scope, estimated case load, and turnaround time.
- Adapt the Laboratory Information System (LIS) to work seamlessly with digital pathology workflows.

### Scanner Requirements

- Evaluate different scanning systems, considering technical requirements and integration into the LIS. See abridged example in **Figure 1**.
- Ensure that scanners meet the intended purpose, including capacity, slide compatibility, and CE-IVD certification for diagnostic purposes.
- Consider scanner maintenance costs and their impact on workflow.

### Output Formats

- Identify ideal scanning profile settings for consistently high picture quality.
- Define file formats for storage and sharing, with a preference for open, non-proprietary formats.
- Specify image size, format, and archiving periods.

### Scanner Validation Study

- Define the scope of the validation study, including tissue sources, stains, and acceptance criteria for diagnostic purposes.
- Establish concordance levels and define severity for non-concordance.
- Create a validation protocol and test a representative sample for each application (e.g., see **Figure 2**).
- Generate a report summarizing the validation aim, results, conclusions, technical requirements, scanner settings, and training evaluations.

### Working Group 2 – Integration of WSI-scanners and DP systems into the Pathology Laboratory Information system

Assuming a validated scanner is in place, these recommendations emphasize the framework for effectively integrating WSI scanners and DP systems into a pathology laboratory information system, ensuring optimal visualization, data management, and workflow efficiency. The recommendations are summarized as follows, with individual statements and agreement levels provided in Appendix B.

**Figure 1.**
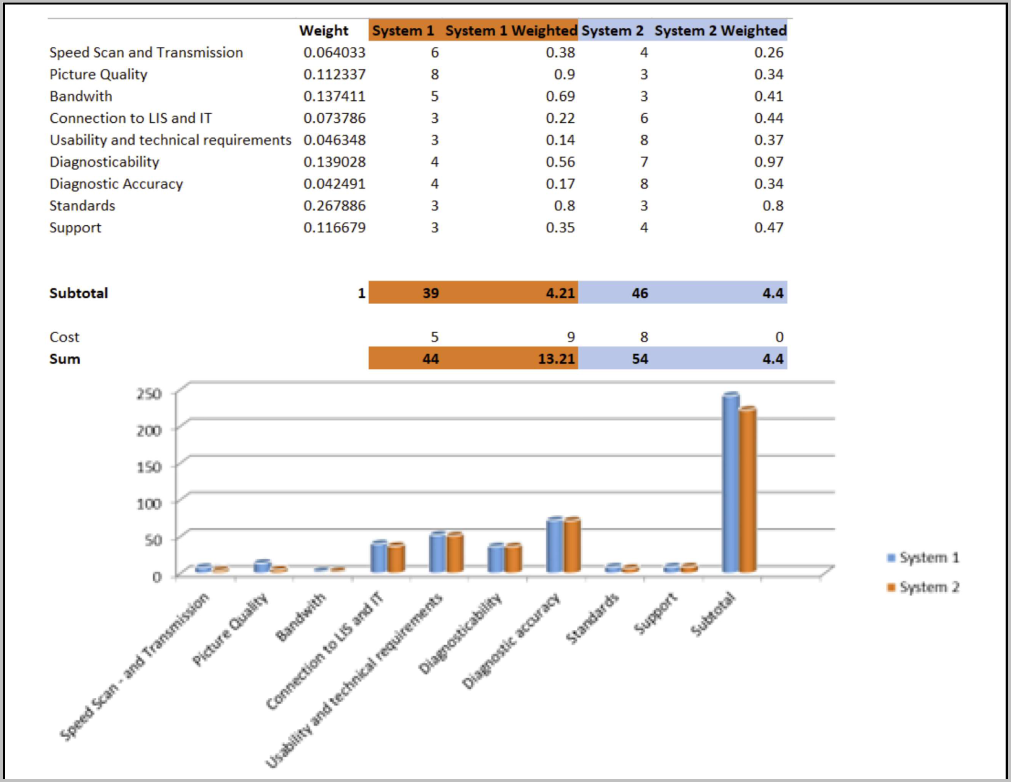
Example of selection criteria compared in two different systems using a scoring model: The criteria should have the same scale (e.g., 1-10) and can be weighted to give more importance to e.g., diagnostic and workflow aspects.

**Figure 2.**
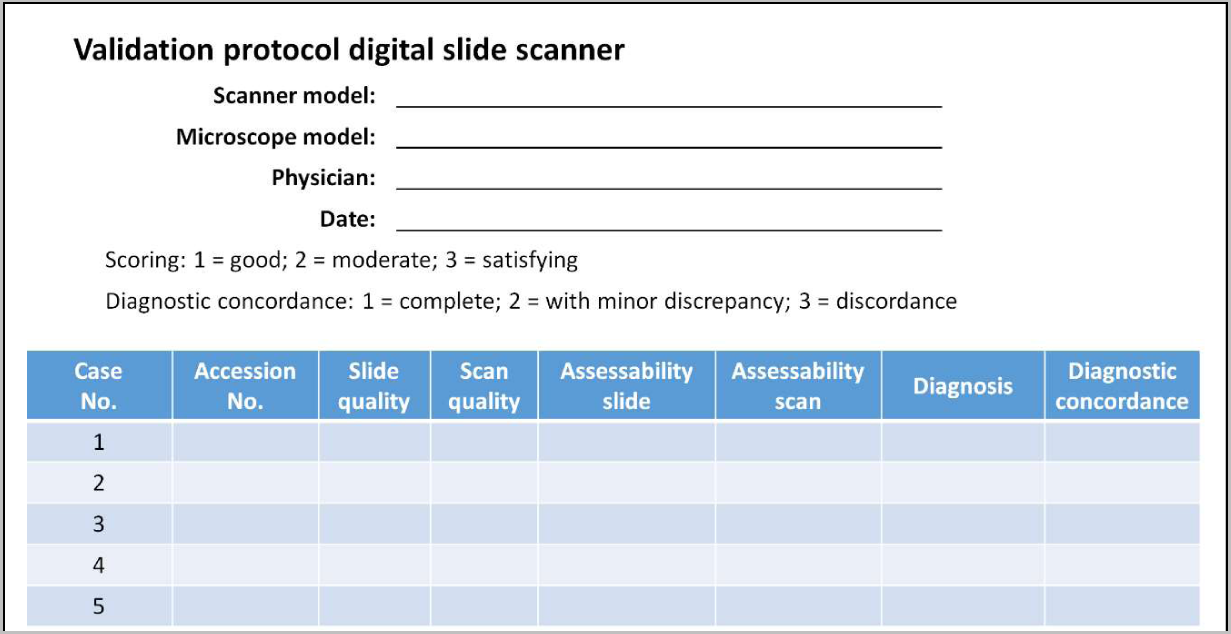
Example validation protocol for comparing diagnosis from glass slide with those of digital slides.

### Visualization (Monitors)

- Larger, high-resolution displays are preferred for better image quality.
- Monitors should be validated by experts and chosen by pathologists.
- Consider ergonomic factors when selecting monitor size.
- Monitor calibration with low deviation is recommended, with documentation.
- Minimum contrast ratio and brightness levels for readability in different lighting conditions are specified.
- Adequate color depth and smooth navigation within viewer software are essential.

### Integration of WSI scanner into Pathology Laboratory Information System (Patho-LIS) for Routine Diagnostics

- The scan workflow should be integrated into the Patho-LIS, Image Management System (IMS), and an image archive, supporting standard communication formats.
- Image data in open formats should be stored in a storage system for retrieval using appropriate streaming mechanisms.
- A secondary test environment is recommended for testing modifications to digital workflow (e.g., new tools).
- Interfaces between WSI scanner and Patho-LIS, as well as between Patho-LIS and IMS, must be established and validated.
- Barcode and alphanumeric codes should be printed on slides for identification.
- Communication protocols should be documented.
- Quality control should be in place before scans are handed over to experts.
- Compliance with research regulations is required for non-routine diagnostic slides.

### Recommendations for IT Interfaces, Standards, and Workflow

- An integrated image viewer should communicate with Patho-LIS and the digital archive.
- The IMS should retrieve necessary information from Patho-LIS to link scanned slides.
- Virtual microscopes should support a comparison view.
- Network speed should meet specific requirements, with a general recommendation of 1 Gbps per scanner.
- High-performance storage solutions should be integrated.
- Redundant installations and alternative workflows are recommended to handle hardware or software malfunctions.

### Working Group 3 – Digital Workflow – compliance with general quality guidelines

These statements are geared towards achieving conformity with current accreditation norms and traceable quality parameters that can be documented within the quality management systems (QMS) of each Institute. The legal framework in which DP enters the stage consists of many facets, from adopted European regulations like in vitro diagnostic regulation (IVDR), to general data protection regulation (GDPR), national legislation like human research act (HRA) and human research law (HRL) and the medical/pathological guidelines of SSPath.

Swiss laboratories regularly perform accreditation^23^ according to the ISO15189^24^ and ISO17025^25^. Relevant elements in terms of quality documentation comprise organizational, procedural, technical, and personnel aspects. For accreditation, DP is regarded as equivalent to conventional histology, and thus tends to benefit from conventional quality control improvements (e.g., usage of barcoding).

In the future, even more improvements via additional quality measurements can be expected with the deployment of DP. For instance, histological sectioning for DP needs more attention by technical personnel, in terms of correct thickness and avoidance of folds, scratches, or peripheral placement of the tissue. Contribution to round robin tests (Quality in Pathology (Germany), European Society of Pathology (ESP), NordiQC) can be fulfilled via slide upload and in-depth calibration measurements. In consequence, DP appears situated to facilitate creation of new more precise standards. The recommendations are summarized as follows, with the individual statements and agreement levels presented Appendix C.

### Quality Requirements for Laboratory Staff and Technicians

- Technicians should receive specific training for digital pathology, including avoiding specimen placement at slide edges and recognizing artifacts.
- Training with scanners and high-tech equipment is recommended for advanced users.
- All workflow steps should be documented in the quality management system’s SOP.
- Consistent barcoding and readable information should be placed on vials, FFPE blocks, slides, and reports.
- Compatibility with additional barcoding solutions for special stainers should be ensured.
- Ordered stainings should quickly appear as placeholders in the digital pathology system.
- The preparation process should be defined to encompass triage of stainings and prioritize highly urgent cases.
- Processes should be defined to switch to regular microscopy for non-digitally compatible microscopy techniques or selection purposes.
- A process should be in place to allow for immediate retrieval of glass slides for rescanning or non-virtual microscopy.
- Emergency plans for severe system errors should be in place.
- Back-up systems for individual components in case of service or maintenance are recommended.
- A process for re-scanning should be in place and counts of re-scanning may serve as a performance test of the scanning process.
- Deviations and problems should be reported within a quality management, or critical incidence reporting, system.

### Additional Quality Requirements for Digital Workflow *Pathologists*

- All pathologists should receive specific training for the digital pathology system, including case management, ordering re-scans, and measurements.
- General knowledge in digital pathology and its potential pitfalls/limitations should be incorporated into the validation process and basic training for the Swiss federal title of pathology.
- Thumbnail images must be compared to scanned images to ensure complete tissue recognition.
- Additional support systems may be included, e.g., tracking systems for hovered areas, annotations for teaching and discussion, and time spent on details. The use of these data should be institutionally regulated and consented by the employed pathologists.
- A digital process for requesting re-scanning should be in place.
- Automated tools like scripts or algorithms should be used cautiously and follow indications, validation, plausibility checks, and quality control.

### IT Support

- IT personnel familiar with the complete system must be in place (in-house or as a service).

### Tumor Boards

- Case presentation at tumor boards can be performed at lower resolutions and in a representative way, but amendments to diagnosis should take place in the diagnostic workstation setting.

### Inter-Institute Tele-Consulting

- The sending institute requesting digital tele-consulting is responsible for slide selection, scanning, resolution, and representativity, with approval declared in the consulting order.
- The receiving pathologist should ensure the diagnosis is made in an appropriate digital setup.
- The institute performing the tele-consulting should document in its sign-out report the number of electronic slides evaluated, viewing platform, and date of access.
- Pathologists may retain digital copies of regions of interest used for consultation.
- Receiving tele-consulting institutes in Switzerland are recommended to validate their workflows within regular accreditation processes.
- To outline the obligations of the asking institute, the sentence “this diagnosis was established on digitized whole slide images kindly provided by the Institute XXX” may be included.
- The final diagnosis and legal liabilities are determined according to the SSPATH guidelines for consultation cases.

### Compliance with Quality Management Systems

- The validation test for the established end-to-end workflow should be documented within the Quality Management system and repeated after major equipment changes.
- SOPs should include all major components of the workflow.
- A re-validation of the complete digital workflow must be performed if major components are replaced.
- Other minor changes due to the modularization of the workflow are handled according to institutional QM guidelines.
- Separate validations must be performed for specific physical measurements.
- DP workflows are expected to increase patient safety and quality measurements, which could be covered with higher reimbursement rates.
- Financial sustainability negotiated with reimbursement agencies should include the needed personnel and equipment under the new DP conditions.

### Working Group 4 – Image analysis (IA)/artificial intelligence (AI)

These statements aim to provide recommendations for the testing, implementation, and quality control of digital image analysis solutions in DP practice.

As part of this process, the working group attempted to concretely define specific terms that are employed within the statements:

1. <u>Image analysis (IA):</u> the extraction of meaningful information from images by means of image processing techniques.
2. <u>Artificial intelligence (AI):</u> the theory and development of computer systems able to perform tasks normally requiring human intelligence, such as visual perception, speech recognition, decision-making, and translation between languages.
3. <u>Machine Learning (ML):</u> the use and development of computer systems that are able to learn and adapt without following explicit instructions, by using algorithms and statistical models to analyze and draw inferences from patterns in data.
4. <u>Deep Learning (DL):</u> a type of machine learning based on artificial neural networks in which multiple layers of processing are used to extract progressively higher-level features from data.
5. <u>Levels of Autonomy</u>: appreciation that unique challenges and requirements are likely needed on a per tasks/algorithms basis depending on the supervision required to safely employ them (see **Table 1**)

**Table 1.** Levels of automation considered by this working group.

| 0 | 1 | 2 | 3 | 4 | 5 |
| --- | --- | --- | --- | --- | --- |
| No automation | Single Task Automation (CAD, Quantification, Prioritization) | Multi Task Automation (Detection + Reporting) | Conditional automation (Only ROI reviewed by specialist) | Full automation in certain conditions (e.g. System finalizes negative prostate biopsies) | Full automation |
| Human Monitors Evaluation |  |  | Automated Systems Monitors Evaluation |  |  |
| Table 1. Levels of automation considered by this working group. |  |  |  |  |  |

To scope the statements, it was discussed that the current state of technology was not sufficiently high to justify consideration of Type 5 algorithms, with type 4 only being considered in niche roles. As such, in the statements below, AI solutions were thought to be aimed to automate repetitive and time-consuming tasks (e.g., mitosis counting, immunohistochemical (IHC)-marker evaluation) and provide a decision support system to the pathologists (e.g., for ambiguous or rare cases). It was noted that the field is very rapidly evolving, and as such, attention should be paid to determine when revision of these statements in light of new inventions, experience, and wisdom is required. The recommendations are summarized as follows, with the individual statements and agreement levels presented in Appendix D.

### General Considerations

- For bioimage analyses and AI-assisted solutions intended for diagnostic use, institutes of pathology should use officially certified systems (e.g., IVD-CE certified, FDA-approved) or laboratory-developed systems that meet validation and quality control requirements.
- The final diagnosis is the responsibility of the pathologist.
- As the level of autonomy in AI systems rises, the interpretability of results becomes more critical.
- Algorithms indicating germline or somatic mutation status must comply with existing laws for molecular testing.
- All systems must fulfill Swiss regulatory requirements.
- AI results must be reported to and reviewed by a board-certified pathologist, following the “integrative diagnosis” paradigm^26^.

### Implementation and Validation of IA/AI Solutions

- Each Institute of Pathology must internally validate IA/AI solutions, even if officially certified systems are used. The scope of validation should be clearly defined.
- Validation should be appropriate for the intended clinical use and clinical setting of the application.
- Validation should involve specimen preparation types relevant to the intended use.
- The validation study should closely emulate the real-world clinical environment.
- Metadata associated with whole slide image (WSI) creation should be documented.
- Diagnoses made using IA/AI should include a version number associated with the validation protocol.
- Revalidation is required whenever a significant change is made to any component of the WSI workflow.
- Known edge cases where IA/AI may not perform well should be documented.
- The pathology report should contain information about the use and regulatory status of IA/AI tools.
- Model performance of on-site validation studies may be included.
- All tissue on a glass slide should be available for computational analysis.
- Quality control measures should ensure the quality of digital images for analysis.
- User requirements and IT requirements for software operation should be clearly defined.
- ROI selection methodology should be stated and described in the diagnostic report.
- The validation process should include a representative set of slides for the intended application.
- Clear descriptions of quality control measures and validation steps should be provided.
- Reproducibility measures, such as pathologist-algorithm correlation, should be documented.
- A validation study should establish diagnostic concordance between digital and glass slides.
- Non-inferiority testing should be carried out between algorithm and pathologists.

### Desirable Technical Properties

- Integration into the existing digital pathology workstation environment is recommended.
- The IA/AI system’s performance must scale with the increasing number of cases.
- Algorithms should highlight regions on digitized slides used to determine their output.
- The ability to provide feedback and prioritize cases or slides is suggested.
- IA/AI can be employed to prioritize cases within work lists or slides.
- Indications should be provided regarding the status of running algorithms.
- Results should be stored in a secure and retrievable manner, in accordance with legal requirements.
- Expected input/output formats should be documented to ensure long-term usability without vendor-specific software.

### Maintenance

- A clear SOP should be in place for the management of hardware and software malfunctions.
- A clear SOP should be in place for the management of updates, including documentation and re-validation requirements.
- The burden of update frequency should be weighed against potential benefits and re-validation costs with awareness of expected algorithm update frequency.

## Conclusion

Using a Delphi process, the members of the Swiss Digital Pathology Consortium reached consensus on practical recommendations for the implementation and validation of digital pathology in clinical workflows. These recommendations focused on its safe usage, with attempts at maximizing patient safety and benefit while minimizing overhead. As a result, we put forward these statements as best-practices to be considered when adopting DP within Switzerland, while also providing another resource for our international colleagues. We are happy to report significant concordance between existing national recommendations and our own, likely due to the converging nature of what appears to be emerging best practices for DP. These recommendations integrate and update upon previous guidelines, providing a dedicated section on the implementation of AI and IA. This fills a niche absent from other recommendations, likely due to the nascent nature of AI/IA field during their creation. Of particular note was that working groups appreciated how rapidly the field is maturing, and realized that, unlike other more established technologies, these DP recommendations will likely need to undergo revisions as technology and the associated implications of this paradigm-shifting technology become more clear.

## Compliance with Ethical Standards

Not applicable

## Funding

Not applicable

## Conflict of Interest

Dr. Janowczyk provides consulting for Merck, Lunaphore, and Roche, the latter of which he also has a sponsored research agreement. Dr. Koelzer has acted as an invited speaker for Sharing Progress in Cancer Care (SPCC) and Indica Labs and holds sponsored research agreements with Roche and IAG. Mr. Kupferschmid is employed by Basys Data GmbH.

## Contributions

AJ, IZ, RG designed the study and edited AJ’s drafted manuscript. All authors (AJ, IZ, SB, VH, MT, JK, TM, DM, MK, RG, TR, LM, IE, GC, KM, VK, DS, WJ, MR, MH, RG) were engaged in conducting the study, drafting and reviewing the statements, and reviewing and approving the final manuscript.

## Data Availability

All data produced in the present study are available upon reasonable request to the authors.

## Appendix A

### Working Group 1 – Scanners, Quality Assurance and validation of Scans Specific statements and agreement levels

Participant Demographics: Statements were voted on by 26 SDiPath members, the composition of which consisted of 73% (19) Pathologists | 15% (4) Researcher | 4% (1) Industry Business Development | 8% (2) Lab Staff/Procurement/IT.

These participants state their place of work to be: 61% (16) University Institute | 23% (6) Public Hospital Institute | 8% (2) Private Institute | 8% (2) Industry.

1 Scope of the diagnostic workflow of Digital Pathology
  1.1 Workflow determination
    1.1.1 Know your scope: The institute should clearly define the scope of the targeted DP-workflow (i.e., intended use). For example: diagnostic biopsy workflow, special stain workflow, image analysis workflow, fluorescence workflow, organ-specific workflow. – 77% (20) Strongly Agree | 23% (6) Agree
    1.1.2 Required Workflow analysis: The entire workflow needs to be inclusive of all workflow components and should consider current and future requirements for long-term flexibility (e.g., paperless transitions). – 73% (19) Strongly Agree | 27% (7) Agree
    1.1.3 The entire workflow needs to be described in a well-documented standard operating procedure (SOP) including its components, the operation and handling of the scanning step, the scanning mode (or modes) for different settings as evaluated in the validation, and the internal quality control (e.g., scan quality, others). – 62% (16) Strongly Agree | 27% (7) Agree | 8% (2) Neutral | 4% (1) Disagree
    1.1.4 SOPs should be written in line with on-site quality management systems and/or accreditation requirements and targets. – 58% (15) Strongly Agree | 38% (10) Agree | 4% (1) Neutral
  1.2 Participants and access authorization
    1.2.1 Security settings of authorized access need to be determined for each component of the workflow, including physical and digital modalities. – 46% (12) Agree | 42% (11) Strongly Agree | 12% (3) Neutral
    1.2.2 Stakeholders (staff, pathologists, technicians, IT) involved in the validation/workflow process need to be informed about their task and receive associated training. – 81% (21) Strongly Agree | 19% (5) Agree
  1.3 Workflow adjustments
    1.3.1 Prioritize workflows based on scope using estimated case load and turnaround time (e.g., all biopsies should be processed first by 11am to ensure same-day sign-out). – 58% (15) Strongly Agree | 27% (7) Agree | 8% (2) Neutral | 8% (2) Disagree
    1.3.2 Define the turnaround time by time recording of the individual work steps, but especially additional work steps: – 42% (11) Agree | 38% (10) Strongly Agree | 19% (5) Neutral
    1.3.3 The Laboratory Information System (LIS) should be adapted to work with digital pathology workflows in terms of clinical information, case management, worklists of attributed cases with the goal of eliminating mismatching of slides from different patients. – 65% (17) Strongly Agree | 35% (9) Agree
2. Scanner requirements
  2.1 Scanner selection criteria
    2.1.1 Evaluate different scanning systems in a pre-selection process for example using a scoring model. Besides technical requirements, integration into the LIS are important items to be considered. – 54% (14) Strongly Agree | 42% (11) Agree | 4% (1) Neutral
  2.2 Technical requirements, to be considered:
    2.2.1 Scanner must meet the intended purpose / must reflect the conditions that are suitable for the intended purpose (including capacity (how many slides/time unit), slide type, scanner slide magazine compatibility, others). – 65% (17) Strongly Agree | 35% (9) Agree
    2.2.2 For diagnostic purposes in pathology, scanner must be CE-IVD certified. – 37% (11) Strongly Agree | 27% (8) Agree | 23% (7) Neutral | 10% (3) Disagree | 3% (1) Strongly Disagree
    2.2.3 Scanner maintenance should be taken into account: costs, effort, effect on workflow (time, frequency). – 50% (13) Strongly Agree | 42% (11) Agree | 8% (2) Neutral
3. Output formats (Refer additionally to WG2)
  3.1 Identify the ideal scanning profile settings associated with the specific workflow, resulting in consistently high picture quality. – 73% (19) Strongly Agree | 23% (6) Agree | 4% (1) Disagree
  3.2 Define file formats for storage, sharing etc. – 62% (16) Strongly Agree | 35% (9) Agree | 4% (1) Neutral
  3.3 Define picture size, format and archiving period. – 54% (14) Strongly Agree | 42% (11) Agree | 4% (1) Neutral
4. Scanner Validation study
  4.1 Validation aim
    4.1.1 Define the scope of the validation study (see WG1-1.1.1) – 65% (17) Strongly Agree | 23% (6) Agree | 12% (3) Neutral
    4.1.2 Select tissue source, stains and techniques (e.g., HE, fluorescence, frozen). – 65% (17) Strongly Agree | 27% (7) Agree | 8% (2) Neutral
    4.1.3 Define the acceptance criteria for diagnostic purpose. – 62% (16) Strongly Agree | 38% (10) Agree
    4.1.4 Define needed concordance level (e.g., same observer >95%). – 58% (15) Strongly Agree | 27% (7) Agree | 15% (4) Neutral
    4.1.5 Define severity for non-concordance (minor-major). – 50% (13) Strongly Agree | 27% (7) Agree | 23% (6) Neutral
    4.1.6 Define deviation (e.g., any finding identified by one modality but not with the other (DP vs mic) with clinical significance. – 54% (14) Strongly Agree | 31% (8) Agree | 15% (4) Neutral
  4.2 Validation protocol and sample size
    4.2.1 Establish a validation protocol based on the validation criteria and test a sufficiently representative sample for each application. – 46% (12) Strongly Agree | 42% (11) Agree | 12% (3) Neutral
    4.3.1 A report summarizing the validation aim, the results, and the final conclusions that points out for what clinical use the DP workflow has been validated and approved. – 62% (16) Strongly Agree | 38% (10) Agree
    4.3.2 The report or the amendments shall also mention the technical requirements, scanner settings, and trainings that were evaluated and determined. – 62% (16) Strongly Agree | 35% (9) Agree | 4% (1) Neutral

## Appendix B

### Working Group 2 – Integration of WSI-scanners and DP systems into the Pathology Laboratory Information system. Specific statements and agreement levels

Participant Demographics: These statements were voted on by 25 SDiPath members, the composition of which consisted of 72% (18) Pathologists | 16% (4) Researcher | 12% (3) Lab Staff/ Procurement/ IT.

These participants state their place of work to be: 60% (15) University Institute | 32% (8) Public Hospital Institute | 4% (1) Private Institute | 4% (1) Industry.

5. Visualization (Monitors)
  5.1 General considerations
    5.1.2 Larger, high-resolution displays show more of the slide at 1:1 magnification (1 screen pixel = 1 image pixel). Lower resolution displays require more panning of the image in order to cover the same physical area. The monitor should be validated by an expert and selected by the pathologist. – 68% (17) Strongly Agree | 20% (5) Agree | 12% (3) Neutral
    5.1.3 When selecting the monitor size, the working distance between the monitor and the pathologist must also be taken into account so that ergonomic working (according to the guidelines of *the Caisse nationale suisse d’assurance en cas d’accidents / Schweizerische Unfallversicherungsanstalt /Istituto nazionale svizzero di assicurazione contro gli infortuni*^1^) is possible. – 68% (17) Strongly Agree | 20% (5) Agree | 12% (3) Neutral
  5.2 Recommendations
    5.2.1 The monitor should have a color calibration option with a low deviation. Automatic self-calibration and adaptation to ambient light is recommended. Manual calibration should be performed at time intervals according to the manufacturer’s recommendations. Calibration steps should be documented. – 44% (11) Strongly Agree | 44% (11) Agree | 8% (2) Neutral | 4% (1) Disagree
  5.3 Brightness and contrast
    5.3.1 The screen should have a minimum contrast ratio of 1000:1 and a brightness of at least 260 cd (candela)/m^2^ in order to maintain high readability in brighter ambient lighting situations. The minimum brightness should be displayable at 0.5 cd/m^2^ or greater^27^. – 44% (11) Strongly Agree | 36% (9) Agree | 20% (5) Neutral
  5.4 Color depth
    5.4.1 The displayable color space should support 24-bit color (8-bit RGB) and 8-bit grayscale. Color depth: Coverage of at least 98% of the Adobe RGB color space is most likely beneficial to display WSI colors accurately^28^. – 48% (12) Strongly Agree | 40% (10) Agree | 12% (3) Neutral
    5.4.2 Navigation devices should allow a smooth and ergonomic slide navigation within the viewer software. – 76% (19) Strongly Agree | 20% (5) Agree | 4% (1) Neutral
6. Integration of the WSI scanner into the pathology information system (Patho-LIS) for routine diagnostics
  6.1 The scan workflow should be integrated into a pathology laboratory information system (Patho-LIS), image management system (IMS) and an image archive. Therefore, it ideally supports standard communication formats such as HL7 and open image format standards (e.g., DICOM). – 80% (20) Strongly Agree | 16% (4) Agree | 4% (1) Neutral
  6.2 All image data in open-formats should optionally be sent to a storage system (e.g., Picture Archiving and Communication System (PACS), Vendor Neutral Archive (VNA)) and retrieved from there using an appropriate streaming mechanism. – 68% (17) Strongly Agree | 24% (6) Agree | 8% (2) Neutral
  6.3 A secondary test environment is recommended to test the respective parameterization of the digital workflow. This system has to function independently from the production system (used for diagnostic) and allows for testing new functionalities, software updates or functional integrations. – 48% (12) Strongly Agree | 32% (8) Agree | 20% (5) Neutral
  6.4 Interfaces between WSI scanner and Patho-LIS or between Patho-LIS and IMS must be established. The application of the interface must be tested by at least one pathologist as part of the required validation study. – 72% (18) Strongly Agree | 28% (7) Agree
  6.5 In addition to the barcode on the glass slides, the alphanumeric code (i.e., sample id and staining) of the slide should also be printed. In this way, the slide can be identified by comparing the recognized barcode with the alphanumeric label and manually corrected, for example, by comparing it with the original slides. – 72% (18) Strongly Agree | 24% (6) Agree | 4% (1) Neutral
  6.6 Any communication protocols should be documented in order to facilitate finding integration issues, ensuring reproducibility and future updates. – 56% (14) Strongly Agree | 44% (11) Agree
  6.7 The scanning process should be verified after hardware or software modifications (e.g., updates, upgrades) in addition to the controls required by national guidelines. – 72% (18) Strongly Agree | 16% (4) Agree | 12% (3) Neutral
  6.8 A quality control step should be in place before the scans are handed over to a medical expert. This can be automated, or by hand. – 64% (16) Strongly Agree | 28% (7) Agree | 8% (2) Neutral
  6.9 Slides used outside of clinical routine diagnostics (e.g., research projects) MUST comply with the Federal Human Research Act (HRA) / *Humanes Forschungsgesetz (HFG) / Loi relative à la recherche sur l’être humain (LRH) / Legge sulla ricerca umana (LRU)*^29^. – 64% (16) Strongly Agree | 20% (5) Agree | 16% (4) Neutral
7. Recommendations for IT interfaces, standards and workflow
  7.1 The image viewer comprised of a) virtual microscope which displays the whole slide images (WSI) of scanned histological sections and b) macroscopic specimen image viewer should be integrated into an image management system (IMS) that allows a (bi)directional communication with Pathology laboratory information system (Patho-LIS) and digital archive. – 72% (18) Strongly Agree | 24% (6) Agree | 4% (1) Neutral
  7.2 If an IMS is used, it must retrieve or pull all necessary information from the Patho-LIS to identify and link the scanned slides to the corresponding LIS entries. – 64% (16) Strongly Agree | 24% (6) Agree | 12% (3) Neutral
  7.3 Virtual microscopes should support a comparison view between photographed (overview images) and scanned images to check WSI for completeness. – 72% (18) Strongly Agree | 24% (6) Agree | 4% (1) Neutral
  7.4 The network speed required for a smooth workflow depends on various parameters (e.g. number of scanners used simultaneously, distance of the scanners to the server, etc.) and must be adjusted according to the specific condition. As a general recommendation, the minimum network speed should be 1 Gbps for each individual scanner. – 72% (18) Strongly Agree | 16% (4) Neutral | 12% (3) Agree
  7.5 The scanner(s) should be connected to a high-performance storage solution (low latency, scale-out architecture, fast transfer speed) that can be integrated into the existing system landscape. – 68% (17) Strongly Agree | 24% (6) Agree | 8% (2) Neutral
  7.7 For the implementation of digital pathology in routine diagnostic, it is recommended to configure the system in such a way that redundant installations (e.g., not only one but at least 2 scanners) and/or an alternative workflow are defined (e.g., maintain the possibility to dispatch the slides) in case of a hardware/software malfunction. – 76% (19) Strongly Agree | 24% (6) Agree
8. Recommendations for archiving
  8.1 To ensure a smooth workflow in DP and daily routine practice, it is recommended to store the WSI for at least 3 months on a high-performance server architecture. – 72% (18) Strongly Agree | 20% (5) Agree | 8% (2) Neutral
  8.2 As a possible currently viable approach archiving of WSI, e.g., in a Picture Archiving and Communication System (PACS)/Vendor Neutral Archive (VNA) for 3 years can be envisaged as a cost-benefit compromise. Such duration of digital archiving would cover the majority of situations in the routine diagnostic workflow where cases need to be compared with previous biopsies. Long-term storage of glass slides should remain unchanged. – 57% (17) Agree | 23% (7) Strongly Agree | 10% (3) Neutral | 10% (3) Disagree
  8.3 The storage concept should take into account compression methods up to the end of the visualization chain, extensive error redundancy during storage, automatic progressive arrangement of the compressed data streams and the patent-free nature of the storage format. – 40% (10) Strongly Agree | 40% (10) Agree | 20% (5) Neutral
  8.4 The acquisition of systems that use industry standards for communication (e.g., DICOM, HL7, CDA, FHIR^2^) with third-party systems (e.g., the hospital information system) or whose systems meet the Integrating the Healthcare Enterprise (IHE) conformance criteria should be preferentially considered for both interoperability and longer-term sustainability (i.e., less likely to become outdated), especially in the environment of larger institutions (e.g., universities/public hospitals). – 48% (12) Strongly Agree | 32% (8) Agree | 20% (5) Neutral

## Appendix C

### Working Group 3 – Digital Workflow – compliance with general quality guidelines Specific statements and agreement levels

Participant Demographics: These statements were voted on by 22 SDiPath members, the composition of which consisted of 73% (16) Pathologists | 18% (4) Researcher | 9% (2) Lab Staff/ Procurement/ IT.

These participants state their place of work to be: 59% (13) University Institute | 27% (6) Public Hospital Institute | 9% (2) Private Institute | 5% (1) Industry.

9. Quality Requirements
  9.1 For laboratory staff and technicians
    9.1.1 All technicians should be specifically trained for digital pathology requirements, e.g., avoid specimen placement at the edge of slides, sensitivity to artefacts like scratches, folds etc. – 77% (17) Strongly Agree | 14% (3) Neutral | 9% (2) Agree
    9.1.2 Training with scanners and other high-tech equipment is recommended to advanced users – 68% (15) Strongly Agree | 18% (4) Agree | 9% (2) Disagree | 5% (1) Neutral
    9.1.3 All workflow steps should be documented in SOP of the quality management system – 82% (18) Strongly Agree | 18% (4) Agree
    9.1.4 Consistent barcoding and readable information should be placed on vials, FFPE blocks, slides and reports. – 82% (18) Strongly Agree | 18% (4) Agree
    9.1.5 Compatibility with additional barcoding solutions for e.g. special stainers should be ensured. – 68% (15) Strongly Agree | 27% (6) Agree | 5% (1) Neutral
    9.1.6 Ordered stainings should quickly appear as space-holders in the DP system. Additional tools to highlight completed cases are recommended. – 59% (13) Strongly Agree | 41% (9) Agree
    9.1.7 Preparation process should be defined taking into consideration existing regular workflows to encompass triage of stainings (Hematoxylin & Eosin, vs. special stains), prolonged drying times, prioritization of highly urgent cases. – 64% (14) Strongly Agree | 32% (7) Agree | 5% (1) Neutral
    9.1.8 Processes should be defined to switch to regular microscopy e.g for not compatible microscopy techniques (polarization) or selection purposes (molecular pathology). – 73% (16) Strongly Agree | 27% (6) Agree
    9.1.9 A process should be in place to allow for immediate retrieval of glass slides for rescanning or non-virtual microscopy. – 82% (18) Strongly Agree | 18% (4) Agree
    9.1.10 Emergency plans for severe system errors should be in place. – 86% (19) Strongly Agree | 14% (3) Agree
    9.1.11 Back-up systems for individual components in case of services/maintenance are recommended. – 82% (18) Strongly Agree | 18% (4) Agree
    9.1.12 Process for re-scanning should be in place. Counts of re-scanning may serve as performance test of the scanning process. – 68% (15) Strongly Agree | 32% (7) Agree
    9.1.13 Deviations and problems should be announced within a quality management or critical incidence reporting system. – 73% (16) Strongly Agree | 27% (6) Agree
  9.2 For digital workflow pathologists
    9.2.1 All pathologists should be specifically trained for the specific DP system, e.g., case management, ordering of re-scans, measurements. – 68% (15) Strongly Agree | 23% (5) Agree | 5% (1) Strongly Disagree | 5% (1) Neutral
    9.2.2 General knowledge in DP and its potential pitfalls/limitations should be incorporated into the validation process (e.g., some cytological features, recognition of microbiota, and those provided by SDiPath, ESP, USCAP, ASCP) and should be part of the basic training for the Swiss federal title of pathology, represented by the *Institut Suisse Pour la Formation Médicale* (ISFM/ SIWF). – 59% (13) Strongly Agree | 27% (6) Agree | 14% (3) Neutral
    9.2.3 All workflow steps should be documented in SOPs of the quality management system. – 82% (18) Strongly Agree | 9% (2) Agree | 9% (2) Neutral
    9.2.4 Thumbnail images must be compared to scanned images to ensure complete tissue recognition. – 77% (17) Strongly Agree | 18% (4) Agree | 5% (1) Neutral
    9.2.5 Additional support systems may be included, e.g. tracking systems for hovered areas, annotations for teaching and discussion, time spent on details. A permanent storage of these data is not mandatory. The use of these data should be institutionally regulated and consented by the employed pathologists. – 50% (11) Strongly Agree | 45% (10) Agree | 5% (1) Disagree
    9.2.6 Ordered stainings should appear as space-holders in the IMS system, to indicate pending stainings. – 77% (17) Strongly Agree | 18% (4) Agree | 5% (1) Neutral
    9.2.7 Processes should be defined to switch to regular microscopy e.g for not compatible microscopy techniques (polarization) or selection purposes (molecular pathology). – 68% (15) Strongly Agree | 32% (7) Agree
    9.2.8 Emergency plans for severe system errors should be in place. – 77% (17) Strongly Agree | 23% (5) Agree
    9.2.9 Digital process for requesting re-scanning should be in place. – 64% (14) Strongly Agree | 32% (7) Agree | 5% (1) Neutral
    9.2.10 Deviations and problems should be announced within a quality management or critical incidence reporting system. – 68% (15) Strongly Agree | 32% (7) Agree
    9.2.11 DP itself has evolved as microscope equivalent in terms of good clinical practice. However, automated tools like scripts or algorithms fall under full considerations of indication, validation, plausibility check and quality control. – 55% (12) Strongly Agree | 36% (8) Agree | 9% (2) Neutral
  9.3 IT-support
    9.3.1 IT-personnel familiar with the complete system must be in place (in house or as service). – 86% (19) Strongly Agree | 14% (3) Agree
  9.4 Tumor boards
    9.4.1 Case presentation at the tumor board can be performed at lower resolutions and in a representative way. However, amendments of diagnosis should take place in the diagnostic workstation setting. – 45% (10) Strongly Agree | 45% (10) Agree | 5% (1) Disagree | 5% (1) Neutral
  9.5 Inter-Institute Tele-consulting
    9.5.1 The sending institute asking for digital tele-consulting is responsible for slide selection, scanning, resolution and representativity. Approval of these steps should be declared in the order for consulting. – 64% (14) Strongly Agree | 36% (8) Agree
    9.5.2 The receiving pathologist should be advised to ensure the diagnosis is made in an appropriate digital setup. – 63% (19) Strongly Agree | 30% (9) Agree | 7% (2) Neutral
    9.5.3 The institute performing the tele-consulting should document in its sign-out report at minimum the number of electronic slides evaluated, viewing platform, and date of access. – 64% (14) Strongly Agree | 32% (7) Agree | 5% (1) Neutral
    9.5.4 Pathologists may want to retain digital copies of the region of interest used for consultation. – 59% (13) Strongly Agree | 36% (8) Agree | 5% (1) Neutral
    9.5.5 Receiving tele-consulting institutes in Switzerland are recommended to validate their workflows within regular accreditation processes – 53% (16) Agree | 43% (13) Strongly Agree | 3% (1) Neutral
    9.5.6 To transparently outline the obligations of the asking institute the sentence “this diagnosis was established on digitized whole slide images kindly provided by the Institute XXX” may be included. – 50% (11) Strongly Agree | 41% (9) Agree | 5% (1) Disagree | 5% (1) Neutral
    9.5.7 Comparable to physical slide sharing, the final diagnosis and legal liabilities are determined according to the SSPATH guidelines for consultation cases (for link to guidelines see below^3^). – 73% (16) Strongly Agree | 27% (6) Agree
    9.5.8 Transmission and access to healthcare information must be ensured according to data protection laws, e.g. HIN secure emails, local servers with controlled access. The responsibility remains with the providing Institute until a national platform is created. – 86% (19) Strongly Agree | 14% (3) Agree
  9.6 Compliance with quality management systems
    9.6.1 The validation test for the established end to end workflow (derived from WP1) should be documented within the Quality Management system and repeated after major equipment changes. – 47% (14) Agree | 47% (14) Strongly Agree | 7% (2) Neutral
    9.6.2 SOPs should include all major components of the workflow – equipment, software versions, scanner models etc. – 73% (16) Strongly Agree | 23% (5) Agree | 5% (1) Neutral
    9.6.3 A re-validation of the complete digital workflow must be performed, if major components are replaced, in particular, a change in scanners, PACS-system, Pathology-LIMS/IMS, software-interfaces etc. – 55% (12) Strongly Agree | 36% (8) Agree | 5% (1) Disagree | 5% (1) Neutral
    9.6.4 Other minor changes due to the modularization of the workflow are handled according to the institutional QM guidelines. This includes new versions of software and equipment without major changes that are integrated with a non-inferior test and simple validation, higher monitor resolution, better graphic power of workstation, faster server conditions, updated scanner same series etc. – 45% (10) Agree | 41% (9) Strongly Agree | 9% (2) Neutral | 5% (1) Disagree
    9.6.5 Separate validations must be performed for specific physical measurements like length or areas as replacement for high power fields. – 45% (10) Strongly Agree | 41% (9) Agree | 14% (3) Neutral
    9.7.1 DP workflows will increase patient safety and quality measurements. The additional technical processes could be covered with higher reimbursement rates as negotiated by SSPATH. – 50% (11) Strongly Agree | 36% (8) Agree | 14% (3) Neutral
    9.7.2 Financial sustainability negotiated with reimbursement agencies should include the needed personnel and equipment under the new DP conditions – 63% (19) Strongly Agree | 30% (9) Agree | 3% (1) Disagree | 3% (1) Neutral

## Appendix D

### Working Group 4 – Image analysis (IA)/artificial intelligence (AI) Specific statements and agreement levels

Participant demographics: These statements were voted on by 24 SDiPath members, the composition of which consisted of 75% (18) Pathologists | 17% (4) Researcher | 8% (2) Lab Staff/ Procurement/ IT.

These participants state their place of work to be: 58% (14) University Institute | 29% (7) Public Hospital Institute | 8% (2) Private Institute | 4% (1) Industry.

10. General considerations
  10.1 For bioimage analyses and AI assisted solutions intended for diagnostic use, Institutes of Pathology should use officially certified (e.g. IVD-CE certified, FDA-approved) systems in the intended manner. Laboratory developed systems may be used as long as the proper validation, quality control, and quality assurance requirements are fulfilled to deliver accurate, precise, and reproducible results. – 47% (14) Agree | 40% (12) Strongly Agree | 13% (4) Neutral
  10.2 The final diagnosis is determined by the pathologist, who is fully legally responsible for it. – 83% (20) Strongly Agree | 12% (3) Agree | 4% (1) Neutral
  10.3 As the level of autonomy rises, interpretability and understanding of how the system is coming to a conclusion becomes more critical. – 58% (14) Strongly Agree | 33% (8) Agree | 8% (2) Neutral
  10.4 Algorithms which may indicate germline or somatic mutation status must follow existing laws for molecular testing. – 71% (17) Strongly Agree | 21% (5) Agree | 4% (1) Neutral | 4% (1) Disagree
  10.5 All these systems must fulfill the Swiss regulatory requirements (e.g. *Heilmittelgesetz*, HMG; *Medizinprodukteverordnung*, MepV; *Verordnung über in-vitro-Diagnostika*, IvDV). – 53% (16) Strongly Agree | 43% (13) Agree | 3% (1) Neutral
  10.6 AI results must be reported to and reviewed by a board-certified pathologist to align with the “integrative diagnosis” paradigm. Unreviewed reporting to third parties should be regarded as off-label use. – 53% (16) Strongly Agree | 40% (12) Agree | 7% (2) Neutral
11. Implementation and validation of IA/AI solutions in diagnostic routine
  11.1 Each Institute of Pathology has to validate the IA/AI solutions internally, even if they are using officially certified systems (e.g., IVD-CE certified, FDA-approved). The scope of the validation should be clearly stated (input / output). – 62% (15) Strongly Agree | 29% (7) Agree | 8% (2) Neutral
    11.1.1 Validation should be appropriate for, and applicable, to the intended clinical use and clinical setting of the application in which the IA/AI algorithms will be employed. The category of algorithm should be described in context with the clinical use (e.g., diagnostic, prognostic, predictive), although it is acknowledged that this will not always be unequivocally possible. – 60% (18) Agree | 27% (8) Strongly Agree | 13% (4) Neutral
    11.1.2 Validation of IA/AI systems should involve specimen preparation types relevant to the intended use (e.g., formalin-fixed paraffin-embedded tissue, frozen tissue, immunohistochemical stains, fluorescence, cytology slides, hematology blood smears). – 71% (17) Strongly Agree | 29% (7) Agree
    11.1.3 The validation study of the IA/AI algorithm system should closely emulate and encompass the real-world clinical environment in which the technology will be used. – 71% (17) Strongly Agree | 25% (6) Agree | 4% (1) Neutral
    11.1.4 Documenting metadata associated with WSI creation must take place, including software versions, firmware versions, understanding that small changes may have large impacts on IA/AI performance. – 62% (15) Strongly Agree | 29% (7) Agree | 8% (2) Neutral
    11.1.5 Diagnoses made using IA/AI should include a version number associated with validation protocol (which contains version numbers of all software/firmware) to enable future contextualization and potential reproduction of results. – 62% (15) Strongly Agree | 33% (8) Agree | 4% (1) Neutral
    11.1.6 Revalidation is required whenever a significant change (e.g., changing of software, firmware updates) is made to any component of the WSI workflow. – 71% (17) Strongly Agree | 29% (7) Agree
    11.1.7 If there are known edge-cases where an IA/AI may not perform well, they should be documented. – 62% (15) Strongly Agree | 38% (9) Agree
  11.2 The pathology report should contain a comment that an IA/AI-tool was used for diagnosis and a comment regarding the regulatory state of the IA/AI solution, e.g. “The tool XY aiding in this diagnosis received a IVD-CE mark in 2022” or “The tool XY aiding in this diagnosis is not currently CE-regulated.” – 53% (16) Strongly Agree | 40% (12) Agree | 7% (2) Neutral
  11.3 Furthermore, model performance of the on-site validation study may be included. – 46% (11) Agree | 38% (9) Strongly Agree | 17% (4) Neutral
  11.4 All tissue that is present on a glass slide should be available and subject to computational analysis, i.e., one needs a verification step to ensure that all relevant tissue areas have been analyzed (whole tissue or relevant hot-spot areas). – 53% (16) Strongly Agree | 40% (12) Agree | 3% (1) Disagree | 3% (1) Neutral
  11.5 A quality control step will be necessary to ensure that the images being analyzed are of suitable quality, for example, regions of blurriness will impact algorithm performance and thus should be alerted to the user. This should include carefully examining whether faint stains, pen marks, foreign objects, air bubbles during sealing, or damage to the cover slide affected the quality of scanned digital images and whether errors such as misalignment of strips or tiles when image stitching has occurred. – 67% (16) Strongly Agree | 29% (7) Agree | 4% (1) Neutral
  11.6 User requirements should be clearly delineated: What level of expertise is required to operate the software? What extent/duration of staff training will be required to effectively utilize the software? – 58% (14) Strongly Agree | 42% (10) Agree
  11.7 Personnel / IT requirements should be clearly delineated: What additional resources (e.g., technologists, hardware, laboratory footprint, IT infrastructure, etc.,) are required to support and ensure optimal function? – 50% (12) Strongly Agree | 50% (12) Agree
  11.8 Regions of interest (ROI) selection methodology should be clearly stated and described (if applicable) in the diagnostic report and whether analysis was on ROI, hot spots, WSI or was based on a pre-selected sample (e.g. tissue microarray (TMA) spot) in order for it to be reliable and reproducible. Selection of ROI or hot spots can be completely automated, completely manual, or a combination of both. The approaches are subject to alternative potential errors. These approaches are likely to be disease and organ-specific. – 40% (12) Agree | 40% (12) Strongly Agree | 17% (5) Neutral | 3% (1) Disagree
  11.9 The validation process should include a large enough set of slides to be fully representative of the intended application (e.g., H&E-stained sections of fixed tissue, frozen sections, cytology, hematology) that reflects the anticipated spectrum and complexity of specimen types, presentation artifacts and variabilities, along with diagnoses likely to be encountered during routine practice. Be aware of “rare” diseases, tissue alterations and aberrant tissue present and how the system handles them (have they been part of the training cohort?). – 67% (16) Strongly Agree | 25% (6) Agree | 8% (2) Neutral
  11.10 Clear descriptions must be provided of quality control measures and validation steps for every clinical assay where image analysis is used. This should include a careful description of algorithm validation. – 67% (16) Strongly Agree | 29% (7) Agree | 4% (1) Neutral
  11.11 Measures of reproducibility should be documented such as pathologist-algorithm correlation and measures of inter-pathologist variability. – 54% (13) Strongly Agree | 38% (9) Agree | 8% (2) Neutral
  11.12 A validation study should establish diagnostic concordance between digital and glass slides for a single observer (i.e., intra-observer variability). – 58% (14) Strongly Agree | 25% (6) Agree | 17% (4) Neutral
  11.13 Non-inferiority testing should be carried out between algorithm and pathologists typically assigned to that particular workflow, and/or against published statistics. – 50% (12) Strongly Agree | 33% (8) Agree | 17% (4) Neutral
12. Desirable technical properties
  12.1 Integration into the existing digital pathology workstation environment is recommended to avoid task-switching burden. – 67% (16) Strongly Agree | 25% (6) Agree | 8% (2) Neutral
  12.2 The performance of an IA/AI-solution must keep up with the increasing number of cases. Analysis of WSIs right after scanning should be supported for time intensive analyses. Algorithm selection should be automated based on available LIS information (e.g. tissue type, staining). – 50% (12) Strongly Agree | 42% (10) Agree | 8% (2) Neutral
  12.3 Algorithms should highlight the regions on the digitized slides, which were used to determine their output, to enable visual control by the pathologist. – 75% (18) Strongly Agree | 25% (6) Agree
  12.4 The ability to provide feedback, to either mark regions or cases as examples of great successes/failures, is suggested. These will allow for both (a) improvement of algorithms, and (b) testing of subsequent versions on real-world difficult/interesting cases. – 58% (14) Strongly Agree | 38% (9) Agree | 4% (1) Neutral
  12.5 Algorithms can be employed to prioritize cases within work lists or slides within cases (i.e., move those cases or slides that the AI algorithms flagged with positive findings to the top of the worklist so that they are reviewed first). – 57% (17) Agree | 23% (7) Strongly Agree | 17% (5) Neutral | 3% (1) Disagree
  12.6 An indication should be provided to clearly advise when the algorithm has yet to be run, or is still running, and may still return additional results to prevent premature sign-out of cases. – 50% (12) Strongly Agree | 42% (10) Agree | 8% (2) Neutral
  12.7 A documentation of where and how the results are stored should be part of the architecture design. Are they in a secured automatically backed up location? Are the results associated with the image itself or the patient file? The results of the algorithm should be stored in a way that diagnostic decisions can be retraced and in accordance with the legal requirements (e.g., screenshot, images of the critical regions). – 54% (13) Strongly Agree | 46% (11) Agree
  12.8 Documenting expected input/output formats is important to ensure they are in “standard” formats (e.g., DICOM, CSV, XLS, etc) that will be easy to share/re-use over the long term. Avoiding the need of using vendor-specific software to access results. – 54% (13) Strongly Agree | 42% (10) Agree | 4% (1) Neutral
13. Maintenance
  13.1 A clear SOP should be in place for the management of hardware and software malfunctions – 67% (16) Strongly Agree | 29% (7) Agree | 4% (1) Neutral
  13.2 A clear SOP should be in place for the management of updates, including a documentation of what the updates consist of, changes to the algorithm and requirements for re-validation. – 62% (15) Strongly Agree | 33% (8) Agree | 4% (1) Neutral
  13.3 Burden of update frequency should be weighed against potential benefits and cost of re-validation of system. Awareness of expected algorithm update frequency is important. – 54% (13) Strongly Agree | 33% (8) Agree | 12% (3) Neutral

## Footnotes

1 https://www.suva.ch/

2 https://www.e-health-suisse.ch/fileadmin/user_upload/Dokumente/E/overview-ihe-hl7-fhir.pdf

3 https://sgpath.ch/qualitaetssicherung/

## Notes

### Funding Statement

This study did not receive any funding

## References

1. Hanna MG, Reuter VE, Samboy J, England C, Corsale L, Fine SW, Agaram NP, Stamelos E, Yagi Y, Hameed M, Klimstra DS, Sirintrapun SJ. Implementation of Digital Pathology Offers Clinical and Operational Increase in Efficiency and Cost Savings. Archives of Pathology & Laboratory Medicine. 2019 Jun 11;143(12):1545–1555.

2. Griffin J, Treanor D. Digital pathology in clinical use: where are we now and what is holding us back? Histopathology. 2017;70(1):134–145.

3. Baidoshvili A, Bucur A, van Leeuwen J, van der Laak J, Kluin P, van Diest PJ. Evaluating the benefits of digital pathology implementation: Time savings in laboratory logistics. Histopathology. 2018 Jun 20; PMID: 29924891

4. Eccher A, Tos APD, Scarpa A, L’Imperio V, Munari E, Troncone G, Naccarato AG, Seminati D, Pagni F. Cost analysis of archives in the pathology laboratories: from safety to management. Journal of Clinical Pathology [Internet]. BMJ Publishing Group; 2023 Aug 2 [cited 2023 Sep 5]; Available from: https://jcp.bmj.com/content/early/2023/08/02/jcp-2023-209035 PMID: 37532289

5. Peyster EG, Janowczyk A, Swamidoss A, Kethireddy S, Feldman MD, Margulies KB. Computational Analysis of Routine Biopsies Improves Diagnosis and Prediction of Cardiac Allograft Vasculopathy. Circulation. 2022 May 24;145(21):1563–1577. PMCID: PMC9133227

6. Janowczyk A, Madabhushi A. Deep learning for digital pathology image analysis: A comprehensive tutorial with selected use cases. J Pathol Inform [Internet]. 2016 Jul 26 [cited 2017 Aug 7];7. Available from: http://www.ncbi.nlm.nih.gov/pmc/articles/PMC4977982/ PMCID: PMC4977982

7. Ho J, Ahlers SM, Stratman C, Aridor O, Pantanowitz L, Fine JL, Kuzmishin JA, Montalto MC, Parwani AV. Can digital pathology result in cost savings? A financial projection for digital pathology implementation at a large integrated health care organization. J Pathol Inform. 2014;5(1):33. PMCID: PMC4168664

8. Dawson H. Digital pathology – Rising to the challenge. Frontiers in Medicine [Internet]. 2022 [cited 2023 Sep 5];9. Available from: https://www.frontiersin.org/articles/10.3389/fmed.2022.888896

9. Retamero JA, Aneiros-Fernandez J, del Moral RG. Complete Digital Pathology for Routine Histopathology Diagnosis in a Multicenter Hospital Network. Archives of Pathology & Laboratory Medicine. 2019 Jul 11;144(2):221–228.

10. Janowczyk A, Baumhoer D, Dirnhofer S, Grobholz R, Kipar A, de Leval L, Merkler D, Michielin O, Moch H, Perren A, Rottenberg S, Rubbia-Brandt L, Rubin MA, Sempoux C, Tolnay M, Zlobec I, Koelzer VH, Swiss Digital Pathology Consortium (SDiPath). Towards a national strategy for digital pathology in Switzerland. Virchows Arch. 2022 Oct;481(4):647–652. PMCID: PMC9534807

11. Unternaehrer J, Grobholz R, Janowczyk A, Zlobec I. Current opinion, status and future development of digital pathology in Switzerland. Journal of Clinical Pathology [Internet]. 2019 Dec 19 [cited 2020 Jan 31]; Available from: https://jcp.bmj.com/content/early/2019/12/19/jclinpath-2019-206155 PMID: 31857377

12. Koelzer VH, Grobholz R, Zlobec I, Janowczyk A. Update on the current opinion, status and future development of digital pathology in Switzerland in light of COVID-19. Journal of Clinical Pathology. BMJ Publishing Group; 2022 Oct 1;75(10):687–689. PMID: 34518361

13. Guide to Digital Pathology [Internet]. [cited 2023 Mar 9]. Available from: https://www.pathologie.de/fachinfos/nachschlagewerke-handbuchreihe/leitfaeden/leitfaeden-detailansicht/?tx_ttnews%5Btt_news%5D=1480&cHash=ec31a0751fdcf421977056e5aafdd387

14. Guidelines for Digital Microscopy in Anatomical Pathology and Cytology [Internet]. The Royal College of Pathologists of Australasia (RCPA); 2020. Available from: https://www.rcpa.edu.au/library/Practising-Pathology/NCRPQF/Docs/Guidelines-for-Digital-Microscopy-in-Anatomical-Pa

15. Evans AJ, Brown RW, Bui MM, Chlipala EA, Lacchetti C, Milner DA, Pantanowitz L, Parwani AV, Reid K, Riben MW, Reuter VE, Stephens L, Stewart RL, Thomas NE. Validating Whole Slide Imaging Systems for Diagnostic Purposes in Pathology. Arch Pathol Lab Med. 2022 Apr 1;146(4):440–450. PMID: 34003251

16. Williams BJ, Brettle D, Aslam M, Barrett P, Bryson G, Cross S, Snead D, Verrill C, Clarke E, Wright A, Treanor D. Guidance for Remote Reporting of Digital Pathology Slides During Periods of Exceptional Service Pressure: An Emergency Response from the UK Royal College of Pathologists. J Pathol Inform. 2020;11:12. PMCID: PMC7245343

17. Going digital: more than just a scanner! [Internet]. SPECTRUM Pathologie. [cited 2023 Mar 9]. Available from: https://www.medmedia.at/spectrum-pathologie/going-digital-more-than-just-a-scanner/

18. Grobholz R. [Digital pathology: The time has come!]. Pathologe. 2018 May;39(3):228–235. PMID: 29691675

19. Zarella MD, Bowman D, Aeffner F, Farahani N, Xthona A, Absar SF, Parwani A, Bui M, Hartman DJ. A Practical Guide to Whole Slide Imaging: A White Paper From the Digital Pathology Association. Arch Pathol Lab Med. 2019 Feb;143(2):222–234. PMID: 30307746

20. Canadian Association of Pathologists Telepathology Guidelines Committee, Bernard C, Chandrakanth SA, Cornell IS, Dalton J, Evans A, Garcia BM, Godin C, Godlewski M, Jansen GH, Kabani A, Louahlia S, Manning L, Maung R, Moore L, Philley J, Slatnik J, Srigley J, Thibault A, Picard DD, Cracower H, Tetu B. Guidelines from the Canadian Association of Pathologists for establishing a telepathology service for anatomic pathology using whole-slide imaging. J Pathol Inform. 2014;5(1):15. PMCID: PMC4023030

21. Chong Y, Kim DC, Jung CK, Kim Dchul, Song SY, Joo HJ, Yi SY. Recommendations for pathologic practice using digital pathology: consensus report of the Korean Society of Pathologists. J Pathol Transl Med. 2020 Nov;54(6):437–452. PMCID: PMC7674756

22. García-Rojo M. International Clinical Guidelines for the Adoption of Digital Pathology: A Review of Technical Aspects. Pathobiology. 2016;83(2–3):99–109. PMID: 27100834

23. Tzankov A, Tornillo L. Hands-On Experience: Accreditation of Pathology Laboratories according to ISO 15189. PAT. Karger Publishers; 2017;84(3):121–129. PMID: 27923229

24. International Organization for Standardization. ISO 15189:2022 – Medical laboratories — Requirements for quality and competence [Internet]. ISO. [cited 2023 Mar 9]. Available from: https://www.iso.org/standard/76677.html

25. International Organization for Standardization. ISO/IEC 17025:2017 – General requirements for the competence of testing and calibration laboratories [Internet]. ISO. 2021 [cited 2023 Mar 9]. Available from: https://www.iso.org/standard/66912.html

26. Beauchamp NJ, Bryan RN, Bui MM, Krestin GP, McGinty GB, Meltzer CC, Neumaier M. Integrative Diagnostics: The Time Is Now—A Report From the International Society for Strategic Studies in Radiology. Journal of the American College of Radiology. Elsevier; 2023 Apr 1;20(4):455–466. PMID: 36565973

27. International Organization for Standardization. ISO 9241-303:2011 – Ergonomics of human-system interaction — Part 303: Requirements for electronic visual displays [Internet]. ISO. [cited 2023 Jun 1]. Available from: https://www.iso.org/standard/57992.html

28. Display Characteristics and Their Impact on Digital Pathology: A Current Review of Pathologists’ Future “Microscope” – PubMed [Internet]. [cited 2023 Mar 9]. Available from: https://pubmed.ncbi.nlm.nih.gov/33042602/

29. SR 810.30 – Federal Act of 30 September 2011 on Research involving Human Beings (Human Research Act, HRA) [Internet]. [cited 2023 Jun 1]. Available from: https://www.fedlex.admin.ch/eli/cc/2013/617/en

